# Safety and Effectiveness of LC16m8 for Pre-Exposure Prophylaxis against mpox in a High-Risk Population: An Open-Label Randomized Trial

**DOI:** 10.1101/2024.06.06.24308551

**Authors:** Nobumasa Okumura, Eriko Morino, Hidetoshi Nomoto, Mashiho Yanagi, Kozue Takahashi, Haruka Iwasaki, Yukari Uemura, Yosuke Shimizu, Daisuke Mizushima, Kazuaki Fukushima, Ei Kinai, Daisuke Shiojiri, Ichiro Itoda, Yasuhiko Onoe, Yoshitomo Kobori, Fukumi Nakamura, Daisuke Tokita, Wataru Sugiura, Norio Ohmagari, Mugen Ujiie

## Abstract

**Background:** The incidence of mpox cases has surged outside endemic regions since May 2022. However, data regarding the safety and efficacy of the LC16m8 vaccine are limited. This study provided opportunities for LC16m8 pre-exposure prophylaxis to high-risk individuals and conducted a randomized controlled trial to assess the effectiveness of LC16m8 in mpox prevention.

**Methods:** This multicenter, randomized, open-label trial enrolled men and women aged ≥18 with high mpox risk. Participants were randomly assigned 1:1 to early or late vaccination groups, receiving vaccinations approximately 70 days apart. Vaccine effectiveness (VE) against mpox development between early and late vaccinations was the primary endpoint. VE against severe mpox, symptoms, “take” incidence, and adverse events were secondary endpoints.

**Results:** A total of 570 and 565 patients were assigned to the early and late vaccination groups, respectively, and 530 and 476 were vaccinated. The median age was 41 years; 99.7% were male, 89.7% were Japanese, and 34.4% had human immunodeficiency virus (HIV). No mpox cases occurred, precluding VE calculations. The take rate was 90.3% (HIV-infected) and 94.6% (uninfected). Adverse events were observed in 97.2% and 98.2% of patients with and without HIV, respectively. No fatal adverse events were observed. Serious adverse events (SAE) were observed in 0.6% (HIV-infected) and 0.5% (uninfected) of patients. One participant without HIV reported pulmonary embolism and deep vein thrombosis as causally undeniable SAE. Local skin reactions: 96.6% (HIV-infected) and 97.9% (uninfected); systemic reactions: 63.6% (HIV-infected) and 64.2% (uninfected).

**Conclusions:** The effectiveness of LC16m8 in mpox remains inconclusive. However, its use in well-controlled HIV-infected and -uninfected individuals showed no significant safety concerns, suggesting the potential for targeted vaccination strategies in at-risk groups. (Japan Registry of Clinical Trials number, jRCT1031230137.)

## Introduction

Monkeypox virus (MPXV) is a double-stranded DNA virus of the genus Orthopoxvirus, first isolated from cynomolgus monkeys in 1958 at the State Serum Institute in Denmark (1). The MPXV has two genetic clades: clade (previously known as the Central African or Congo Basin clade) and clade (the West African clade). Clade is further divided into two subclades: a and b (2). The number of mpox cases has increased, especially in West and Central Africa, with outbreaks in the Democratic Republic of Congo in 1996–1997 (3) and in Nigeria in 2017–2018 (4).

Since May 2022, the number of mpox cases has risen in countries outside the endemic regions. The global number of patients with mpox peaked in August 2022 and has been declining since then. However, as of February 23, 2024, 93,921 confirmed cases and 179 deaths worldwide have been reported (5). In Japan, the number of patients peaked between March and May 2023. However, the number of patients has since decreased, with 242 cases reported as of March 15, 2024 (6). The route of transmission and patient background differentiate this epidemic from the pre-2022 mpox (7). According to the World Health Organization, 83.3% of mpox cases are sexually transmitted, 85.4% of patients are men who have sex with men (MSM), and 52.0% of patients are individuals with human immunodeficiency virus (HIV) (5). The case fatality rate of mpox in the current epidemic is low at 0.3% (5); however, when limited to people with advanced HIV, the fatality rate increases to 7% and 27% for people with a CD4 cell count of < 350 cells/μL and < 100 cells/μL, respectively (8). Although antiviral drugs such as tecovirimat (9–12) and cidofovir (9, 13) are the most promising candidates for treatment, few studies have investigated their efficacy, and preventing mpox is crucial.

Vaccination is a key preventive strategy for mpox infections. Vaccines against the smallpox virus, which belongs to the same genus Orthopoxvirus as the MPXV, establish cross-immunity to mpox in vaccinees (14). Representative smallpox vaccines include Dryvax (first generation), ACAM2000 (second generation), Modified Vaccinia Ankara (MVA), and LC16m8 (third generation). LC16m8 is an attenuated live vaccine with restricted replication, developed using the LC16m8 strain. This strain was successfully isolated by passaging the Lister strain of the vaccinia virus, which has been used for the smallpox vaccine. LC16m8 has low virulence because it cannot express the full length of B5R owing to a frameshift mutation in the *V5R* gene, which encodes a membrane protein and cannot complete the extracellular enveloped virion stage of the viral life cycle (15). LC16m8 is inoculated singly and intradermally using a bifurcated needle and multiple-puncture technique. A local reaction occurs at the site of inoculation, generally from a few days to two weeks later, with erythema, swelling, then blisters or pustules with a central umbilical fossa, and finally crusting, which is referred to as “take” (16–18), which is a surrogate indicator of immunogenicity (19).

In Japan, approval was granted in 1975 to produce and sell vaccines using the LC16m8 strain; however, because the national program for smallpox vaccination was discontinued in 1976, the LC16m8 vaccine has not been marketed since then. With the outbreak of the mpox epidemic, an urgent need arose to provide LC16m8 immunization to those at high risk. In August 2022, the use of LC16m8 was extended to include the prevention of mpox in addition to smallpox (20). LC16m8 was first used for the post-exposure prophylaxis of mpox (21), and its immunogenicity and safety have been tested in healthy adults (16). The effectiveness of pre-exposure prophylaxis has been reported for MVA in the United Kingdom (22), United States (23–26), Israel (27), and Spain (28); however, the effectiveness and safety have not been studied for LC16m8. Therefore, this study offered opportunities for LC16m8 pre-exposure prophylaxis to high-risk individuals, including patients with HIV, and conducted a randomized controlled trial to evaluate the effectiveness and safety of LC16m8 in preventing mpox.

## Methods

### Ethical considerations

This study adhered to the principles of the Declaration of Helsinki and the Ethical Guidelines for Medical and Biological Research Involving Human Subjects. This study was approved by the Institutional Review Board of the National Center for Global Health and Medicine (NCGM-S-004692-00). All participants provided written informed consent.

### Trial design

A multicenter randomized open-label study was conducted. Participants were recruited and enrolled at eight sites in Tokyo, Japan, between June and October 2023. Eligible participants were randomly assigned in a 1:1 ratio to the early or late vaccination groups. Randomization was conducted online according to a computer-generated randomization code with two allocation factors: year of birth (before 1975 or after 1976) and whether the participant was an MSM. Because the smallpox vaccine was discontinued in Japan in 1976, the birth year was used to determine whether the participant had at least one history of smallpox vaccination (Supplementary Appendix).

### Study population

This study included men and women aged 18 years or older who provided written consent and were at a high risk of mpox infection. High-risk patients were defined as follows: A) patients with HIV who have continuously been receiving anti-HIV therapy and who had a CD4 cell count of > 200 cells/μL within six months before study enrollment, B) those who had a negative HIV screening test in the month before study enrollment and who met any of the following conditions; a) MSM (any sexuality such as gay, bisexual, or transgender) with a history of sexually transmitted bacterial infections (e.g., syphilis, gonorrhea, chlamydial infection, or *Mycoplasma genitalium* infection) within the past year, who had participated in group sex within the past year, or had two or more sexual partners, b) those taking pre-exposure prophylaxis for HIV infection (PrEP).

The exclusion criteria included those 1) with a history of mpox; 2) received the smallpox vaccine since 2022; 3) had at least a moderate contact risk within 14 days with patients with mpox; 4) immunocompromised patients (e.g., those taking immunosuppressive drugs, such as oral corticosteroids, cyclosporine, tacrolimus, and azathioprine, but have been continuously treated with anti-HIV therapy and who have been confirmed to have a CD4 cell count of > 200 cells/μL in the six months before study enrollment did not meet these criteria.), 5) had anaphylaxis owing to the smallpox vaccine components, 6) pregnant or may have been pregnant, or 7) judged to be inappropriate for inclusion in the study by the principal investigator. Moderate contact risk with patients with mpox included a history of contact within 1 m, contact with normal skin or mucous membranes, including wounds, or contact with family members and cohabitants.

### Intervention

All the participants were vaccinated at the National Center for Global Health and Medicine (NCGM). The freeze-dried cell culture smallpox vaccine, LC16 “KMB” (KM Biologics, Kumamoto, Japan), was provided by the Ministry of Health, Labour and Welfare. Approximately 0.01 mL of LC16 after lysis was inoculated into the skin on the lateral side of the upper arm using the multiple-puncture technique with 15 compressions using a bifurcated needle.

The early vaccination group was inoculated on specific days at the time of enrollment, whereas the late vaccination group was inoculated approximately 70 days after the paired participants in the early vaccination group were inoculated. If vaccination was judged inappropriate through an interview on the day of vaccination, the vaccine should be administered to a group assigned to the subsequent inoculation opportunities. The participants were instructed not to change their sexual behavior before and after vaccination.

### Endpoints

The primary endpoint was vaccine effectiveness (VE) against mpox development, which was evaluated by comparing the incidence of mpox between the early and late vaccination groups during the focused observation period. The focused observation period was defined as the period from Day 14 after the day of vaccinating participants in the early vaccination group (Day 0) to the day of vaccinating paired participants in the late vaccination group (Day 70). Participants were instructed to visit their assigned research institution if they developed mpox symptoms during the focused observation period. Research institutions were asked to report the onset of mpox to the research office if patients had been diagnosed with mpox. Mpox diagnosis was made using nucleic acid amplification testing, as is the usual practice.

The secondary endpoints included VE against hospitalization or death due to mpox during the focused observation period (defined as a severe mpox), mpox symptoms, the incidence of take, the incidence of adverse events, and VE against mpox development during the entire study period. The “takes” were judged in two ways: *sensu lato* and *sensu stricto*. *Sensu lato* was defined as the appearance of any gross local inflammatory reaction, such as erythema, swelling, induration, blisters, or crusting at the inoculation site. *Sensu stricto* was defined as the appearance of induration, blistering, or crusting at the inoculation site (29). *Sensu stricto* was further subdivided into two categories: one involving grade 0 or greater induration (i.e., induration of any diameter), blisters, and crusting (Definition 1), and the other involving grade 1 or greater induration (i.e., induration > 2.5 cm), blisters, and crusting (Definition 2) (16, 30). Adverse events were serious if they led to death, were life-threatening, required hospitalization for treatment or prolonged hospitalization, caused permanent or notable impairment or malfunction, or caused a congenital anomaly in the descendants. The principal investigator or co-investigator determined the causal relationship between all adverse events and the study.

### Data collection

The participants maintained diaries and visited the hospital, as scheduled in the Supplementary Appendix. Briefly, on the day of registration (visit 1), 1) demographic characteristics were collected, including date of birth, sex, and race; 2) past medical histories, comorbidities, concomitant medications, and allergies; and 3) mpox risk backgrounds including HIV infection, CD4 cell count in the last six months (only in HIV-positive participants), anti-HIV drugs in use (only in HIV-positive participants), HIV-PrEP (only in HIV-negative participants), sexual orientation, gender identity, history of sexually transmitted bacterial infection in the last year, history of attending group sex in the last year, and having two or more current sexual partners. Visit 2 was on the day of vaccination (Day 0). Visits 3 and 4 corresponded to days 14 and 28, respectively, and information on adverse events was collected at these two time points. The pre-specified adverse events, including local skin reactions (injection site pain, redness, swelling, induration, blisters, crust, and itching), lymphadenopathy, and systemic symptoms (fever, chills, headache, malaise, arthralgia, myalgia, rash at other sites than the injection site, vomiting, and diarrhea) were collected and graded based on patient self-reports, and the other adverse events were collected by free description. All adverse events were graded (Table S1). In the early vaccination group, visit 5 was on Day 70, which corresponded to the vaccination date of the paired participants in the late vaccination group. Visits 3, 4, and 5 were conducted via telephone or online (e-mail or questionnaire). An e-mail was sent to the participants at the end of the study (visit 6 in the early vaccination group and visit 5 in the late vaccination group) to retrospectively confirm that they had not developed mpox during the study period.

### Target sample size

The target number of participants was 5,000 (2,500 in each group). The number of cases required to confirm the hypothesis is 32.5 cases under an enrollment of 5,000 participants, assuming the following conditions: 1:1 allocation ratio, 90% power, a one-sided significance level of 2.5%, expected mpox incidence rate (IR) of 1% during the prevaccination period in the late vaccination group, and VE for mpox development ≥ 70% between the early and late vaccination group.

### Statistical analyses

The Intention to Treat (ITT) analysis population comprised all the allocated participants. The modified ITT (M-ITT) analysis population comprised all participants in the ITT analysis population, excluding those who violated the inclusion or exclusion criteria, developed mpox, or contracted mpox by Day 13 of vaccination, in which all efficacy endpoints were missing, with serious protocol violation during the evaluation of the primary endpoint, or in which vaccination was canceled because of the inappropriate condition for vaccination (the provisions of 6.2.3, Supplementary Appendix) after inclusion in the early vaccination group. The M-ITT2 analysis population comprised all participants in the M-ITT analysis population, excluding those not vaccinated with LC16m8. The safety analysis population comprised all participants who received the LC16m8 vaccination.

The primary VE analysis was performed using the M-ITT population. Supplementary analyses were also performed for the ITT and M-ITT2 populations. The safety analysis population was used to evaluate the incidence of take and adverse events.

Of the data collected as participants’ background, continuous variables were described in summary statistics (mean, standard deviation, median, maximum, and minimum values), and categorical variables were described as the number and percentage.

VE (%) was calculated as (1-relative risk (RR)) ×100. The RR of the IR and its 95% confidence interval (CI) were estimated using Poisson regression with the onset of mpox as the response variable and the treatment group as the explanatory variable. The RR was calculated as (IR in the early vaccination group) / (IR in the late vaccination group). IR was calculated as (number of mpox cases) / (total person-years observed). VE for severe mpox and mpox during the entire study period was estimated in the same manner. For mpox symptoms, the number and proportion of participants in each group and the pooled groups were calculated. Subgroup analyses for VE were conducted for the following variables: sex, year of birth (before 1975 or after 1976), MSM status, HIV infection, and sexually transmitted bacterial infections in the last year.

The number of cases, percentage, and 95% CI were calculated for the incidence of take and adverse events. A secondary analysis for the take incidence was performed by dividing the participants into groups via HIV infection status. Because the take incidence is lower in those previously vaccinated against smallpox than those who have not (17, 31), the year of birth (before 1975 or after 1975) was also included as an explanatory variable, and the adjusted risk ratio for HIV infections was estimated using a risk ratio regression model. Adverse events were analyzed using Fisher’s exact test for the presence of HIV infection. Unless otherwise specified, the P-value was based on a two-sided test with a significance threshold of < 0.05. SAS version 9.4 (SAS Institute, Cary, NC, USA) and R version 4.3.1 (R Foundation for Statistical Computing, Vienna, Austria) were used for the statistical analyses.

## Results

### Participant selection

A total of 1,135 participants were recruited from June 1, 2023, to October 6, 2023; 570 patients were assigned to the early vaccination group, and 565 participants were assigned to the late vaccination group (ITT analysis population), of which 530 and 476 were vaccinated, respectively (safety analysis population) (Figure 1). Subsequently, 568 and 561 patients in the early and late vaccination groups, respectively, were included in the M-ITT population analysis. Demographic characteristics of the M-ITT population are presented in Table 1. The background factors collected from the early and late vaccination groups were well-balanced.

**Figure 1.**
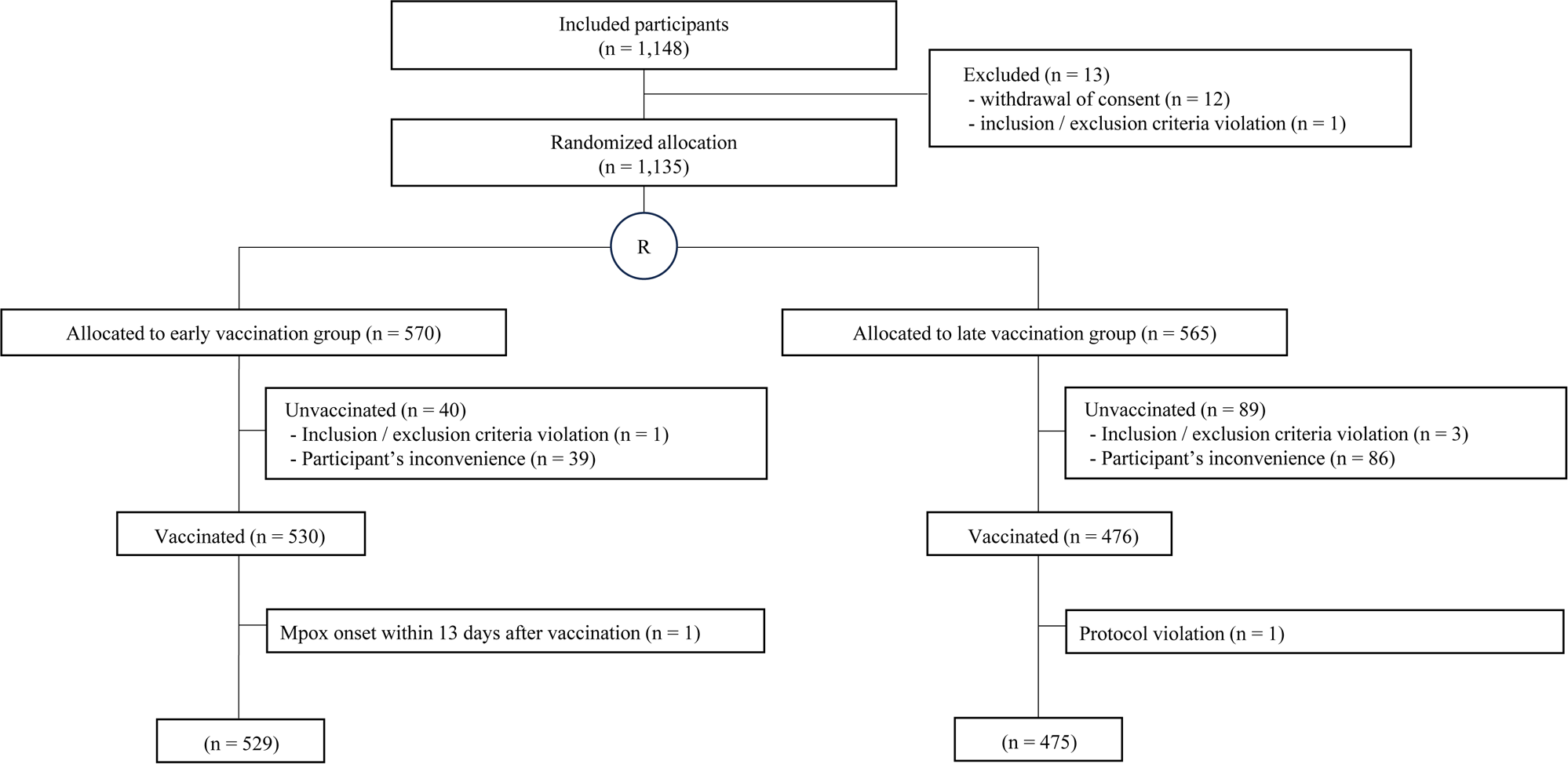
Participant selection flowchart.

**Table 1.**
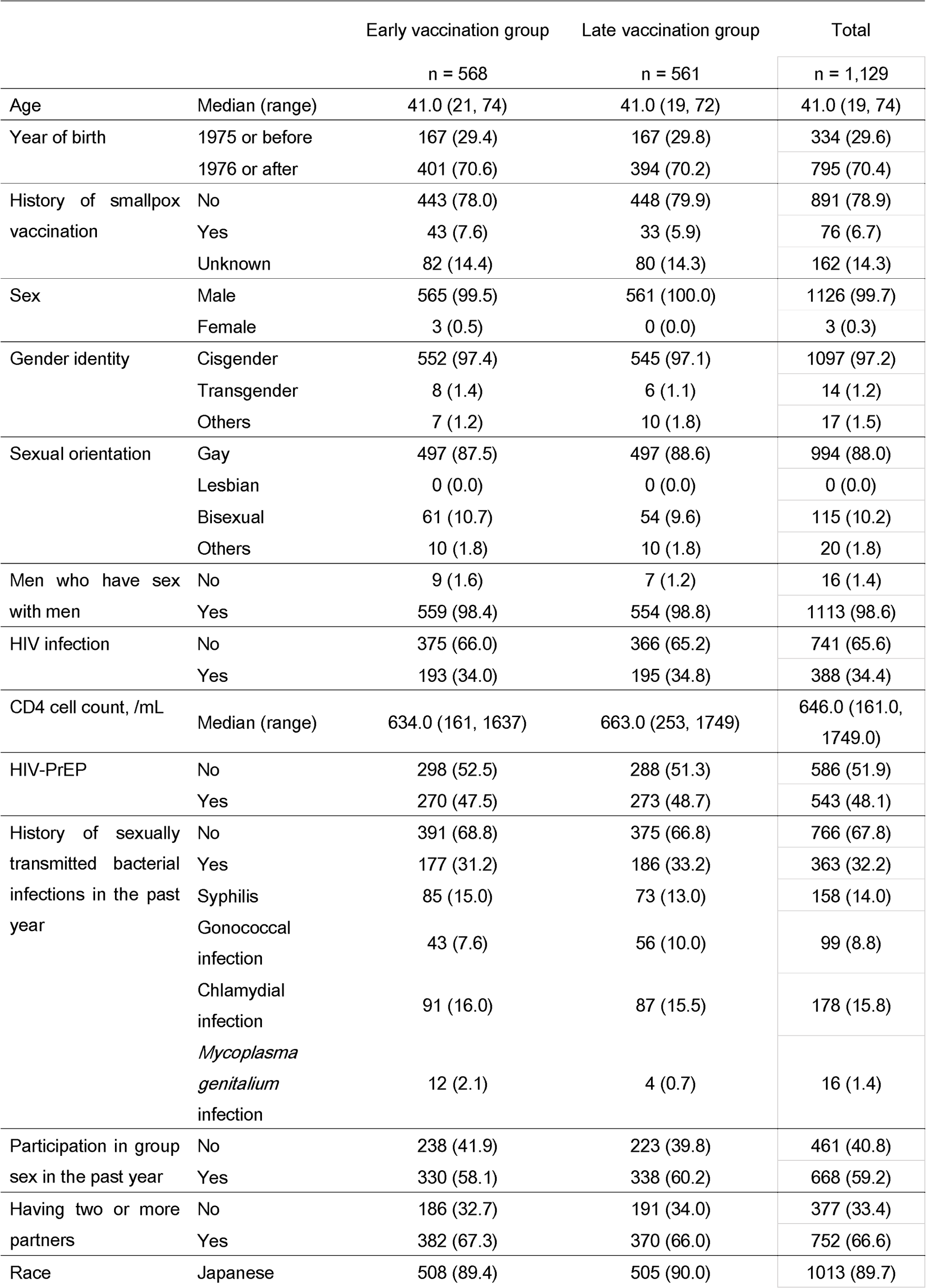
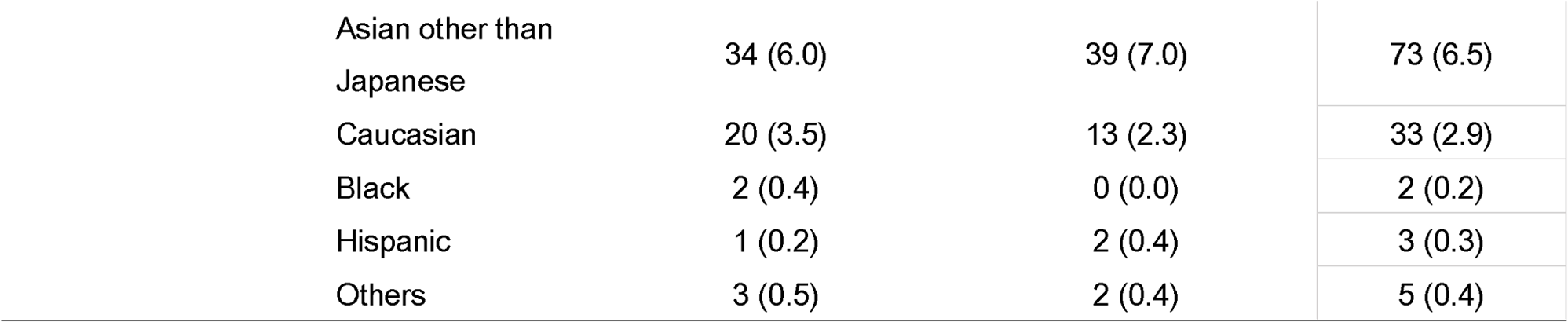
Participants background in the modified Intention to Treat analysis population.

|  |  | Early vaccination group | Late vaccination group | Total |
| --- | --- | --- | --- | --- |
|  |  | n = 568 | n = 561 | n = 1,129 |
| Age | Median (range) | 41.0 (21, 74) | 41.0 (19, 72) | 41.0 (19, 74) |
| Year of birth | 1975 or before | 167 (29.4) | 167 (29.8) | 334 (29.6) |
|  | 1976 or after | 401 (70.6) | 394 (70.2) | 795 (70.4) |
| History of smallpox vaccination | No | 443 (78.0) | 448 (79.9) | 891 (78.9) |
|  | Yes | 43 (7.6) | 33 (5.9) | 76 (6.7) |
|  | Unknown | 82 (14.4) | 80 (14.3) | 162 (14.3) |
| Sex | Male | 565 (99.5) | 561 (100.0) | 1126 (99.7) |
|  | Female | 3 (0.5) | 0 (0.0) | 3 (0.3) |
| Gender identity | Cisgender | 552 (97.4) | 545 (97.1) | 1097 (97.2) |
|  | Transgender | 8 (1.4) | 6 (1.1) | 14 (1.2) |
|  | Others | 7 (1.2) | 10 (1.8) | 17 (1.5) |
| Sexual orientation | Gay | 497 (87.5) | 497 (88.6) | 994 (88.0) |
|  | Lesbian | 0 (0.0) | 0 (0.0) | 0 (0.0) |
|  | Bisexual | 61 (10.7) | 54 (9.6) | 115 (10.2) |
|  | Others | 10 (1.8) | 10 (1.8) | 20 (1.8) |
| Men who have sex with men | No | 9 (1.6) | 7 (1.2) | 16 (1.4) |
|  | Yes | 559 (98.4) | 554 (98.8) | 1113 (98.6) |
| HIV infection | No | 375 (66.0) | 366 (65.2) | 741 (65.6) |
|  | Yes | 193 (34.0) | 195 (34.8) | 388 (34.4) |
| CD4 cell count, /mL | Median (range) | 634.0 (161, 1637) | 663.0 (253, 1749) | 646.0 (161.0, 1749.0) |
| HIV-PrEP | No | 298 (52.5) | 288 (51.3) | 586 (51.9) |
|  | Yes | 270 (47.5) | 273 (48.7) | 543 (48.1) |
| History of sexually transmitted bacterial infections in the past year | No | 391 (68.8) | 375 (66.8) | 766 (67.8) |
|  | Yes | 177 (31.2) | 186 (33.2) | 363 (32.2) |
|  | Syphilis | 85 (15.0) | 73 (13.0) | 158 (14.0) |
|  | Gonococcal infection | 43 (7.6) | 56 (10.0) | 99 (8.8) |
|  | Chlamydial infection | 91 (16.0) | 87 (15.5) | 178 (15.8) |
|  | *Mycoplasma genitalium* infection | 12 (2.1) | 4 (0.7) | 16 (1.4) |
| Participation in group sex in the past year | No | 238 (41.9) | 223 (39.8) | 461 (40.8) |
|  | Yes | 330 (58.1) | 338 (60.2) | 668 (59.2) |
| Having two or more partners | No | 186 (32.7) | 191 (34.0) | 377 (33.4) |
|  | Yes | 382 (67.3) | 370 (66.0) | 752 (66.6) |
| Race | Japanese | 508 (89.4) | 505 (90.0) | 1013 (89.7) |

|  |  |  |  |
| --- | --- | --- | --- |
| Asian other than Japanese | 34 (6.0) | 39 (7.0) | 73 (6.5) |
| Caucasian | 20 (3.5) | 13 (2.3) | 33 (2.9) |
| Black | 2 (0.4) | 0 (0.0) | 2 (0.2) |
| Hispanic | 1 (0.2) | 2 (0.4) | 3 (0.3) |
| Others | 3 (0.5) | 2 (0.4) | 5 (0.4) |

### Demographic characteristics

Demographic characteristics of the M-ITT analysis population are presented in Table 1. The median age of the participants was 41 (range 19-74) years, and 334 (29.6%) were born in or before 1975. Seventy-six (6.7%) participants responded that they had received the smallpox vaccine before this study. Almost all participants (99.7%) were male, and 98.6% were MSM. Regarding race, Japanese accounted for the majority of cases with 1,013 (89.7%), followed by non-Japanese Asians with 73 (6.5%) and Caucasians with 33 (2.9%). A total of 34.4% of the patients were HIV-infected, and the median CD4 cell count was 646 /μL. Of the participants, 32.2% had a history of sexually transmitted bacterial infections within the past year; of which 15.8% had chlamydial infections, 14.0% had syphilis, 8.8% had gonococcal infections, and 1.4% had *M. genitalium* infections. The background factors collected from the early and late vaccination groups were well balanced.

### VE for preventing mpox

The incidence rate of mpox was 0 (0/82.6 person-years) in the early vaccination group and 0 (0/92.0 person-years) in the late vaccination group in the M-ITT analysis population during the focused observation period. The VE of LC16m8 for mpox development could not be calculated (Table 2). No cases of mpox in the ITT and M-ITT2 populations were reported, and VE could not be calculated (Table 2).

**Table 2.** Vaccine effectiveness for mpox development during the focused observation period.

|  | N | Cases | Mpox development |  | Vaccine effectiveness* |  |  |
| --- | --- | --- | --- | --- | --- | --- | --- |
|  |  |  | Observed person-years | Incidence rate<br>(/100 person-years) | VE (%) | 95% CI | *P*-value** |
| M-ITT analysis population |  |  |  |  |  |  |  |
| Early vaccination group | 568 | 0 | 82.554 | 0.00 | - | - | - |
| Late vaccination group | 561 | 0 | 91.978 | 0.00 |  |  |  |
| ITT analysis population |  |  |  |  |  |  |  |
| Early vaccination group | 570 | 0 | 82.710 | 0.00 | - | - | - |
| Late vaccination group | 565 | 0 | 92.183 | 0.00 |  |  |  |
| M-ITT2 analysis population |  |  |  |  |  |  |  |
| Early vaccination group | 529 | 0 | 82.554 | 0.00 | - | - | - |
| Late vaccination group | 475 | 0 | 78.557 | 0.00 |  |  |  |

\*: $VE (\%) = (1-RR) \times 100$ RR (relative risk) estimated by modified Poisson regression. RR is synonymous with incidence rate ratio in this study.
\*\* : one-sided test
M-ITT, modified Intention to Treat
CI, confidence interval
RR, risk ratio
VE, vaccine effectiveness

The total observation period of 1,129 participants in the M-ITT analysis population before and after vaccination was 193.7 and 314.1 person-years, respectively. The onset of mpox was not observed in either period, and VE could not be calculated (Table S2). VE could not be calculated in any subgroup analyses.

### VE for preventing severe mpox

During the focused observation period, the incidence rate of mpox cases requiring hospitalization or death in the M-ITT analysis population was zero in the early and late vaccination groups. The VE could not be calculated (Table S3); therefore, the mpox symptoms could not be evaluated.

### Vaccine take

In the safety analysis population, the proportion of take *sensu lato* was 96.2% (968/1,006), the proportion of take *sensu stricto* (Definition 1) was 93.1% (937/1,006), and the proportion of take *sensu stricto* (Definition 2) was 92.3% (929/1,006) (Table S4).

The proportion of take by HIV status was then compared between the groups. Take *sensu lato* (97.2% vs. 94.3%, P = 0.024), *sensu stricto* (Definition 1) (94.6% vs. 90.3%, P = 0.013), and *sensu stricto* (Definition 2) (93.9% vs. 89.5%, P = 0.018) were significantly higher in HIV-uninfected individuals than in HIV-infected individuals (Table 3). The risk ratios for being born in 1976 or later and having an HIV infection for take *sensu lato* were 0.999 (95% CI: 0.973–1.026, P = 0.965) and 0.970 (95% CI: 0.942–0.999, P = 0.040), respectively. When a similar analysis was conducted for take *sensu stricto* (Definition 1), the risk ratios were 1.039 (95% CI: 0.998–1.082, P = 0.066) and 0.961 (95% CI: 0.924–0.998, P = 0.041), respectively, indicating that HIV infection negatively impacted the proportion of take as much as having a birth year before 1975 (Table 4).

**Table 3.** Take proportion by HIV status.

| HIV |  | Number of cases | Take *sensu lato* |  | Take *sensu stricto*<br>(definition 1) |  | Take *sensu stricto*<br>(definition 2) |  |
| --- | --- | --- | --- | --- | --- | --- | --- | --- |
|  |  |  | n (%) | 95% CI | n (%) | 95% CI | n (%) | 95% CI |
| Infected | Early vaccination group | 185 | 177 (95.7) | 91.7, 98.1 | 168 (90.8) | 85.7, 94.6 | 167 (90.3) | 85.1, 94.1 |
|  | Late vaccination group | 167 | 155 (92.8) | 87.8, 96.2 | 150 (89.8) | 84.2, 94.0 | 148 (88.6) | 82.8, 93.0 |
|  | Total | 352 | 332 (94.3) | 91.4, 96.5 | 318 (90.3) | 86.8, 93.2 | 315 (89.5) | 85.8, 92.5 |
| Uninfected | Early vaccination group | 345 | 340 (98.6) | 96.7, 99.5 | 329 (95.4) | 92.6, 97.3 | 328 (95.1) | 92.2, 97.1 |
|  | Late vaccination group | 309 | 296 (95.8) | 92.9, 97.7 | 290 (93.9) | 90.6, 96.3 | 286 (92.6) | 89.0, 95.2 |
|  | Total | 654 | 636 (97.2) | 95.7, 98.4 | 619 (94.6) | 92.6, 96.2 | 614 (93.9) | 91.8, 95.6 |
| Difference*(95% CI) |  |  | -2.9% (-5.7, -0.2) |  | -4.3% (-7.8, -0.8) |  | -4.4% (-8.1, -0.7) |  |
| P-value |  |  | 0.024 |  | 0.013 |  | 0.018 |  |

\* HIV-infected — HIV-uninfected
※Fisher's exact test
HIV, human immunodeficiency virus
CI, confidence interval

**Table 4.**
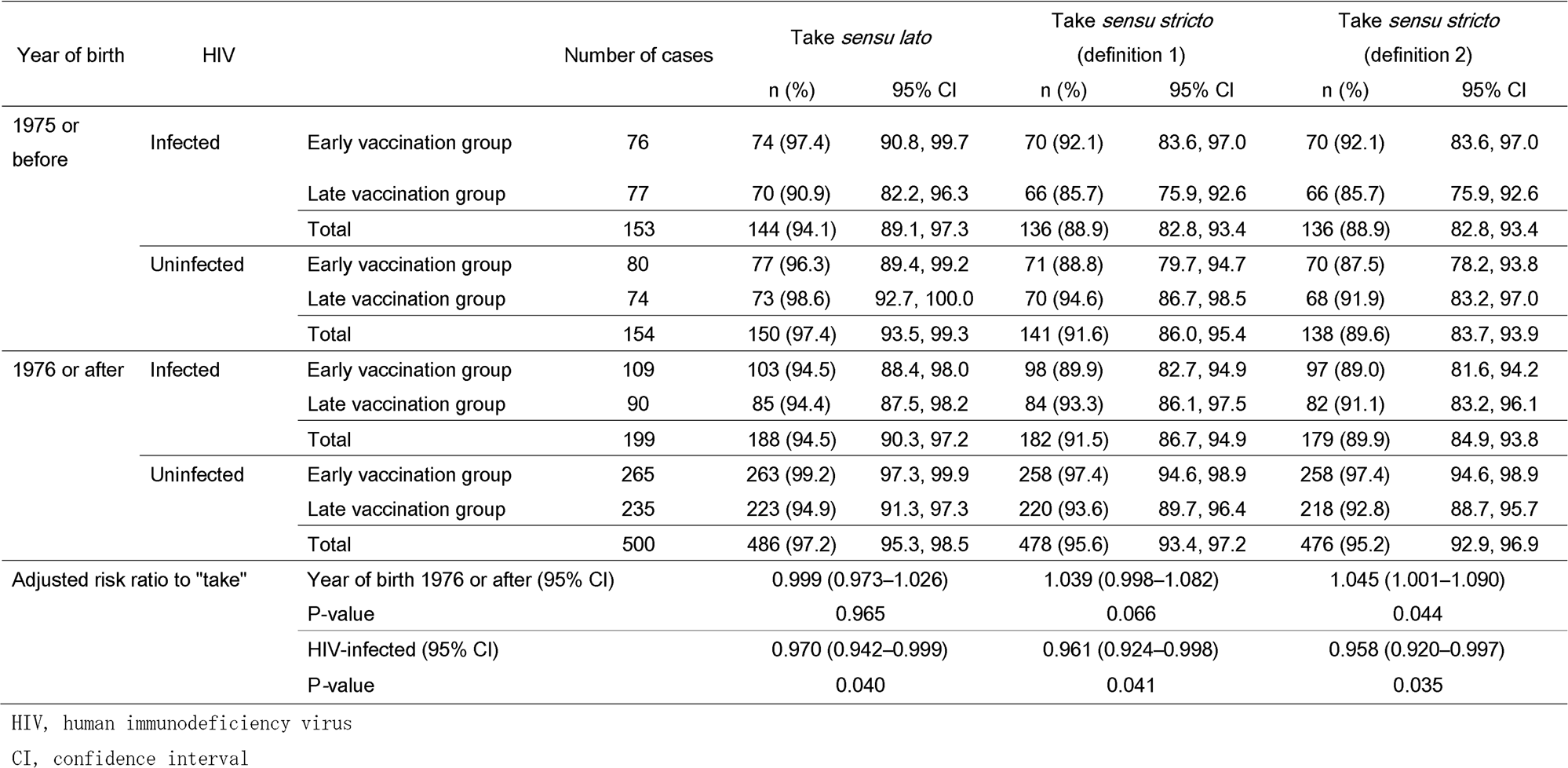
Take proportion by HIV status and year of birth.

### Safety

Adverse events were observed in 97.8% (984/1,006) of the safety analysis population, almost all of which were causally related to vaccination. Adverse events were compared between participants with and without an HIV infection (Table 5). No significant differences were observed between the two groups for any adverse events (97.2% vs. 98.2%, P = 0.366), adverse events causally related to the vaccine (96.9% vs. 98.0%, P = 0.282), pre-specified local skin reactions (96.6% vs. 97.9%, P = 0.297), or pre-specified systemic symptoms (63.6% vs. 64.2%, P = 0.890). When limiting adverse events to grade 3 or greater, patients with HIV had significantly fewer adverse events (15.9% vs. 22.6%, P = 0.014) and pre-specified local skin reactions (6.5% vs. 11.5%, P = 0.014) than those without HIV. No significant difference was observed in pre-specified systemic symptoms between the two groups, even when limited to grade 3 or greater (11.4% vs. 15.1%, P = 0.104). The proportion of patients with axillary lymphadenopathy was significantly lower among patients with HIV (22.2% vs. 33.5%, P < 0.001) than in those without HIV.

**Table 5.** Adverse events.

|  | Patients with HIV<br>n = 352 | Patients without HIV<br>n = 654 | Total<br>n = 1,006 | Risk difference* (95% CI) | P-value** |
| --- | --- | --- | --- | --- | --- |
| All adverse events | 342 (97.2) | 642 (98.2) | 984 (97.8) | -1.0 (-3.0, 1.0) | 0.366 |
| Adverse events (grade 3 or greater) | 56 (15.9) | 148 (22.6) | 204 (20.3) | -6.7 (-11.7, -1.7) | 0.014 |
| Causally related adverse events | 341 (96.9) | 641 (98.0) | 982 (97.6) | -1.1 (-3.2, 1.0) | 0.282 |
| Pre-specified local skin reactions | 340 (96.6) | 640 (97.9) | 980 (97.4) | -1.3 (-3.5, 0.9) | 0.297 |
| Pain | 138 (39.2) | 286 (43.7) | 424 (42.1) | -4.5 (-10.9, 1.8) | 0.181 |
| Redness | 302 (85.8) | 581 (88.8) | 883 (87.8) | -3.0 (-7.4, 1.3) | 0.160 |
| Swelling | 251 (71.3) | 477 (72.9) | 728 (72.4) | -1.6 (-7.5, 4.2) | 0.605 |
| Induration | 142 (40.3) | 277 (42.4) | 419 (41.7) | -2.0 (-8.4, 4.4) | 0.547 |
| Blister | 197 (56.0) | 415 (63.5) | 612 (60.8) | -7.5 (-13.9, -1.1) | 0.021 |
| Crusting | 287 (81.5) | 561 (85.8) | 848 (84.3) | -4.2 (-9.1, 0.6) | 0.084 |
| Itching | 312 (88.6) | 597 (91.3) | 909 (90.4) | -2.6 (-6.6, 1.3) | 0.180 |
| Pre-specified local skin reactions (grade 3 or greater) | 23 (6.5) | 75 (11.5) | 98 (9.7) | -4.9 (-8.5, -1.4) | 0.014 |
| Lymphadenopathy | 91 (25.9) | 229 (35.0) | 320 (31.8) | -9.2 (-15.0, -3.3) | 0.003 |
| Axillary lymphadenopathy | 78 (22.2) | 219 (33.5) | 297 (29.5) | -11.3 (-17.0, -5.7) | <0.001 |
| Pre-specified systemic symptoms | 224 (63.6) | 420 (64.2) | 644 (64.0) | -0.6 (-6.8, 5.6) | 0.890 |
| Fever | 65 (18.5) | 123 (18.8) | 188 (18.7) | -0.3 (-5.4, 4.7) | 0.933 |
| Chills | 51 (14.5) | 118 (18.0) | 169 (16.8) | -3.6 (-8.3, 1.2) | 0.158 |
| Headache | 92 (26.1) | 179 (27.4) | 271 (26.9) | -1.2 (-7.0, 4.5) | 0.710 |
| Malaise | 128 (36.4) | 244 (37.3) | 372 (37.0) | -0.9 (-7.2, 5.3) | 0.784 |
| Arthralgia | 67 (19.0) | 99 (15.1) | 166 (16.5) | 3.9 (-1.0, 8.8) | 0.130 |
| Myalgia | 69 (19.6) | 115 (17.6) | 184 (18.3) | 2.0 (-3.1, 7.1) | 0.442 |
| Rash other than the injection site | 41 (11.6) | 61 (9.3) | 102 (10.1) | 2.3 (-1.7, 6.3) | 0.273 |
| Vomiting | 3 (0.9) | 4 (0.6) | 7 (0.7) | 0.2 (-0.9, 1.4) | 0.701 |
| Diarrhea | 39 (11.1) | 64 (9.8) | 103 (10.2) | 1.3 (-2.7, 5.3) | 0.515 |
| Pre-specified systemic symptoms (grade 3 or greater) | 40 (11.4) | 99 (15.1) | 139 (13.8) | -3.8 (-8.1, 0.5) | 0.104 |
| Serious adverse events | 2 (0.6) | 3 (0.5) | 5 (0.5) | 0.1 (-0.8, 1.1) | 1.00 |
| Adverse events leading to death | 0 (0.0) | 0 (0.0) | 0 (0.0) | 0.0 (0.0, 0.0) | 1.00 |

\* HIV-infected—HIV-uninfected
\*\* Fisher's exact test
HIV, human immunodeficiency virus
CI, confidence interval

Five cases of serious adverse events (SAE) were reported: two in patients with HIV and three in patients without HIV. SAE in the patients with HIV included a left testicular tumor (diffuse large B-cell lymphoma) and amoebic dysentery, whereas SAE in the uninfected participants included bilateral pulmonary embolism (PE)/deep vein thrombosis (DVT), *Campylobacter* gastroenteritis, and condylomata acuminata. The only case in which a causal relationship with vaccination could not be ruled out was bilateral PE/DVT. This patient experienced left lower extremity pain and dyspnea 11 days after LC16m8 vaccination. Ten days later, the patient visited a medical institution and was diagnosed with PE/DVT, which improved with oral apixaban. No deaths occurred during the study.

## Discussion

To our knowledge, this is the first study to examine the effectiveness of pre-exposure vaccination with LC16m8 to prevent mpox. Although live vaccines such as measles, rubella, varicella, and mumps have been considered contraindicated in patients with HIV, they are permissible for patients whose viral load has been suppressed by antiretroviral therapy and whose CD4 cell count is > 200 cells/μL (32). In addition, the smallpox vaccine guidelines state that patients with HIV who have been exposed to the smallpox virus or are at high risk of infection should receive the live vaccine ACAM2000 if their CD4 cell counts are > 200 cells/μL (33). Therefore, this study included HIV-infected individuals as vaccinees for LC16m8 on the condition that they were on antiretroviral therapy and had a CD4 cell count of > 200 cells/μL.

Studies that have evaluated the VE of MVA primarily focused on men at a high risk of mpox (22, 27, 34), whereas other studies also included women (23, 26); a US study using an electronic health record database included approximately 9% women (23). Another US study, using jurisdictional health departments’ case registries, did not include cisgender women and included approximately 1% of transgender men (26). In the present study, women could participate if they were HIV-PrEP users; however, only three women participated. Therefore, almost all the participants in this study were MSM, suggesting there may be fewer female PrEP users in Japan or that women may have a lower sense of crisis regarding mpox even though there are a certain number of female PrEP users.

No mpox cases were observed in the M-ITT analysis population, and the VE could not be calculated. This result may be due to the fact that the incidence rate of mpox in Japan decreased during the study period and the lack of power caused by the inability to include the pre-specified number of participants. Therefore, the incidence of mpox in Tokyo was estimated during the study period as a reference. Because the exact number of HIV-PrEP users in Tokyo is unknown, the number of MSM at risk was estimated. As of January 2023, the male population of Tokyo was 6,797,186, of which 5,044,346 were aged 20–74 (35), corresponding to the age group of the participants in this study. Furthermore, because the percentage of MSM in Japan is approximately 2% of the male population (36), the number of MSM was approximately 100,887. In Japan, mpox is designated as a class IV infectious disease, and doctors who diagnose it are required to report all cases. The number of patients with mpox between June 21 and December 27, 2023, was 41 in Japan and 28 in Tokyo, which included the observation period of this study (37). Based on these values, the estimated cumulative incidence of mpox among MSM in Tokyo is 0.028% (95% CI: 0.018–0.040). Because the denominator, MSM, includes those who are not sexually active, the cumulative incidence of mpox in Tokyo was likely underestimated. Nonetheless, the mpox cumulative incidence in this study was 0% (95% CI: 0.00–0.033), which was lower than the underestimated cumulative incidence in Tokyo.

As MVA effectively prevents mpox in other countries (22–28), the need to provide vaccination opportunities for high-risk populations in Japan using the available LC16m8 vaccine is increasing. Because LC16m8 is pharmaceutically approved for mpox prophylaxis but not available on the market, it can only be used within the framework of a study to examine its efficacy and safety. Having a non-inoculated group in this study was not ethically desirable due to the approval of the vaccine for mpox in Japan. Additionally, it was necessary to complete the vaccination of high-risk individuals who wanted to be vaccinated within a short period.

Before the development of LC16m8, Japanese documents defined take as “complete pox,” which were blisters or pustules with a pox umbilicus (a depression in the center), and “incomplete pox,” which were papules or nodules with infiltration leading to vesicles (38). Similarly, the US Centers for Disease Control and Prevention states that “A normal take will show a pustular lesion or an area of definite induration or congestion surrounding a central lesion, which can be a scab or an ulcer. Anything else is considered a “non-take” (29), which agreed with a previous study (17). In another study, healthy adults were inoculated with LC16m8 to examine immunogenicity. Take in the broad sense was defined as the presence of erythema, swelling, induration, blisters, and crusting, whereas take in the narrow sense was defined as the presence of grade 1 or higher induration, blisters, or crusting. (16). As grade 0 was not assigned to blisters and crusting, grade 0 adverse events were essentially excluded only in induration. Because the definition of take is heterogeneous, this study used multiple definitions to calculate its proportion. Another study reported take proportions of 94.4% and 86.6% for primary vaccinees and revaccinees, respectively (17), which were similar to those of participants born after 1976 and before 1975 in the present study. Additionally, Morino et al. reported that take in the broad sense was observed in 93.8% (45/48) and the narrow sense in 80% (40/50) of the participants in their study (16), both of which were slightly lower than the proportions of *sensu lato* and *sensu stricto* (Definition 2) in this study. A possible reason for this discrepancy is the difference in the number of compressions during inoculation. In the present study, the number of compressions was uniformly set to 15, whereas in the study by Morino et al., it was set to 5 or 10 depending on the history of smallpox vaccination, which may have resulted in takes in more participants in the present study. Another factor to consider is the reporting method. In this study, the presence or absence of take was judged by the researcher based on the participants’ self-report, whereas in the study by Morino et al., the researcher visually checked the participants’ arms to determine the presence or absence of take. In the case of self-reporting, some participants may not have reported local skin reactions due to the time and effort required for reporting, which could have led to an underestimation of the take proportion in the present study. However, because the take proportion was higher in this study, the difference in reporting methods possibly did not affect the difference in take proportion between the two studies.

The present also examined the effects of HIV infection on take. Regardless of the definition of take, patients with HIV had a significantly lower proportion of take than individuals without HIV. The proportion of take is lower among smallpox revaccinees than among primary vaccinees (17); therefore, this study used multivariate analysis using a risk ratio regression model in which the year of birth was also included as an explanatory variable. As in the univariate analysis, HIV infection negatively affected the take proportion; however, the risk ratio was approximately 0.96, which does not have a large clinical impact.

Pre-specified local skin reactions and systemic symptoms were observed in 97.4% and 64.0% of participants, respectively. Pre-specified local skin reactions are generally synonymous with take. Regarding pre-specified systemic symptoms, Saito et al. reported fever (2.0%), headache (0.5%), diarrhea (0.2%), and arthralgia (0.1%) (17), all of which were observed less frequently than in the present study. Morino et al. reported fatigue in 44%, headache in 28%, myalgia in 26%, arthralgia in 22%, and diarrhea in 16% of participants (16), similar to the results of the present study. Axillary lymphadenopathy was identified in approximately 30% of the participants in this study, which is comparable to the 28% reported by Morino et al. but more frequent than the 9.0% reported by Saito et al. The frequency of adverse events differed significantly between the present study and that reported by Saito et al. owing to two possible reasons. The first is the difference in the methods used to collect adverse events. In this study, local and systemic adverse events were defined in advance, and participants were asked to record the presence or absence of these events in their diaries in a timely manner, which may have resulted in fewer forgotten reports. The second reason is the proportion of primary vaccinees with a higher frequency of take and adverse events than revaccinees (17). The proportion of primary vaccinees in the Saito et al. cohort was 47.5%, whereas that in this study (i.e., those born after 1976) was 70.4%, which may have partially contributed to the high adverse events rate.

This study also evaluated the safety of LC16m8 in patients with HIV for the first time. No significant differences were observed in the incidence of overall adverse events, pre-specified local skin reactions, or pre-specified systemic symptoms between patients with and without HIV. The incidence of grade 3 or higher adverse events and axillary lymphadenopathy was significantly lower in patients with HIV than in those without HIV. None of the SAE reported for older-generation smallpox vaccines (39), such as generalized vaccinia, progressive vaccinia, encephalitis, myopericarditis, or vaccinia keratitis, have been reported in patients with HIV. However, one case of PE/DVT was causally related to SAE in this study. Because the patient who developed PE/DVT had a family history of pulmonary thrombosis, a hereditary predisposition to thrombosis was expected; however, blood tests did not confirm this, and a vaccine-related relationship could not be ruled out. However, PE and DVT are also adverse events associated with other vaccines, such as MVA (40, 41) and various COVID-19 vaccines (42), suggesting that these adverse events are not LC16m8-specific. Thus, using LC16m8 in well-controlled patients with and without HIV had no significant safety concerns.

The fact that take and axillary lymphadenopathy occur less frequently in patients with HIV than in patients without HIV is favorable from the viewpoint of adverse events; however, this could also be a sign of inferior immunogenicity in these patients (19). In a previous study examining the immunogenicity and safety of MVA in patients with HIV with a CD4 cell count of > 200 cells/μL, the geometric mean titer of antibodies against the vaccinia virus was significantly higher in healthy participants than in those with HIV; however, the plaque reduction neutralization test geometric mean titer value at 26 weeks post-vaccination follow-up was similar in both groups (43). A smaller study with a CD4 cell count of > 350 cells/μL reached similar conclusions (44). When anti-MPXV-neutralizing antibodies were measured in MVA-inoculated individuals, lower levels of neutralizing antibodies were observed in patients with HIV than in uninfected individuals; however, no significant differences were observed (45). Thus, although these data are from studies using MVA, immunogenicity to vaccinia virus and MPXV in patients with HIV is expected to be equal to or very mildly inferior to that in uninfected individuals. Real-world data analyzing the actual association between immunosuppression or HIV infection and vaccine effectiveness have not shown a significant association (27, 46). Although an accurate evaluation of immunogenicity in patients with HIV receiving LC16m8 is pending, LC16m8 vaccination in these adequately controlled by antiretroviral therapies would be acceptable because no new safety concerns were observed.

This study has several limitations. The first is behavioral bias in evaluating VE. This study used a design in which all participants were vaccinated with LC16m8 with a time lag. Participants might refrain from risky behaviors for mpox up to vaccination and then increase risky behaviors after vaccination, potentially leading to an increase in mpox cases among vaccinees. Therefore, participants were instructed not to change their sexual behavior before and after the LC16m8 vaccination to prevent this. As a result, no mpox cases in either group were observed, eliminating the need to consider the impact of behavioral bias on the study. Second, this study excluded people with HIV with a CD4 cell count of < 200 cells/μL. Although people with advanced HIV infection—who are considered to be at the highest risk of death (8)—should be protected from mpox, LC16m8 administration in patients with HIV with a CD4 cell count of < 200 cells/μL was considered contraindicated because of its replication competence. However, the cocooning strategy should indirectly protect people with advanced HIV by increasing vaccination coverage among those at high risk of mpox living in the same communities.

However, this study also had some strengths. Most overseas studies examining the efficacy of MVA are database-based and retrospective, with potential bias due to uncollected confounding factors. This study used randomization and followed participants prospectively; therefore, there was little concern about bias. In addition, the safety of LC16m8 in pediatric patients has been confirmed by the experience of inoculating approximately 50,000 Japanese children with LC16m8 (47).

In conclusion, the use of LC16m8 in well-controlled individuals with and without HIV showed no significant safety concerns, suggesting the potential for targeted vaccination strategies in at-risk groups. The findings of this study indicate a promising avenue for future research and development of vaccines tailored to specific populations, potentially improving public health outcomes in vulnerable communities. Furthermore, the freeze-dried formulation of LC16m8 offers logistical advantages, and immunity can be achieved with a single immunization. Therefore, LC16m8 could contribute to ending the global mpox epidemic.

## Data Availability

All data produced in the present study are available upon reasonable request to the authors

## Declarations

### Ethics approval

This study adhered to the principles of the Declaration of Helsinki and the Ethical Guidelines for Medical and Biological Research Involving Human Subjects. This study was approved by the Institutional Review Board (NCGM-S-004692-00).

### Author contributions

NOk, EM, HN, YU, YS, DM, and MU conceptualized the study.

NOk, EM, HN, MY, KT, HI, YU, YS, DM, and MU developed the methodology.

DT, WS, NOh and MU were responsible for project administration.

MY, KT, HI, DM, KF, EK, DS, II, YO, YK, and FN recruited participants.

NOk, EM, HN, MY, KT, HI, and MU conducted and supervised the vaccination.

YU and YS did the statistical analyses.

NOk prepared the first draft of the article, which was revised by EM, HN, and MU.

All authors reviewed the manuscript, contributed to its interpretation, and approved the final version.

## Acknowledgments

We thank the participants of the study, the staff at the National Center for Global Health and Medicine, and other research institutions and research team members. We thank the nursing department staff for providing the LC16m8 inoculations and Keiko Tanaka for the appropriate assignment of the nursing department staff. We thank the staff of the Pharmaceutical Department, especially Takahiro Nishimura and Yasukata Ohashi, for the proper storage and dispensing of LC16m8.

---

**Figure S1.**
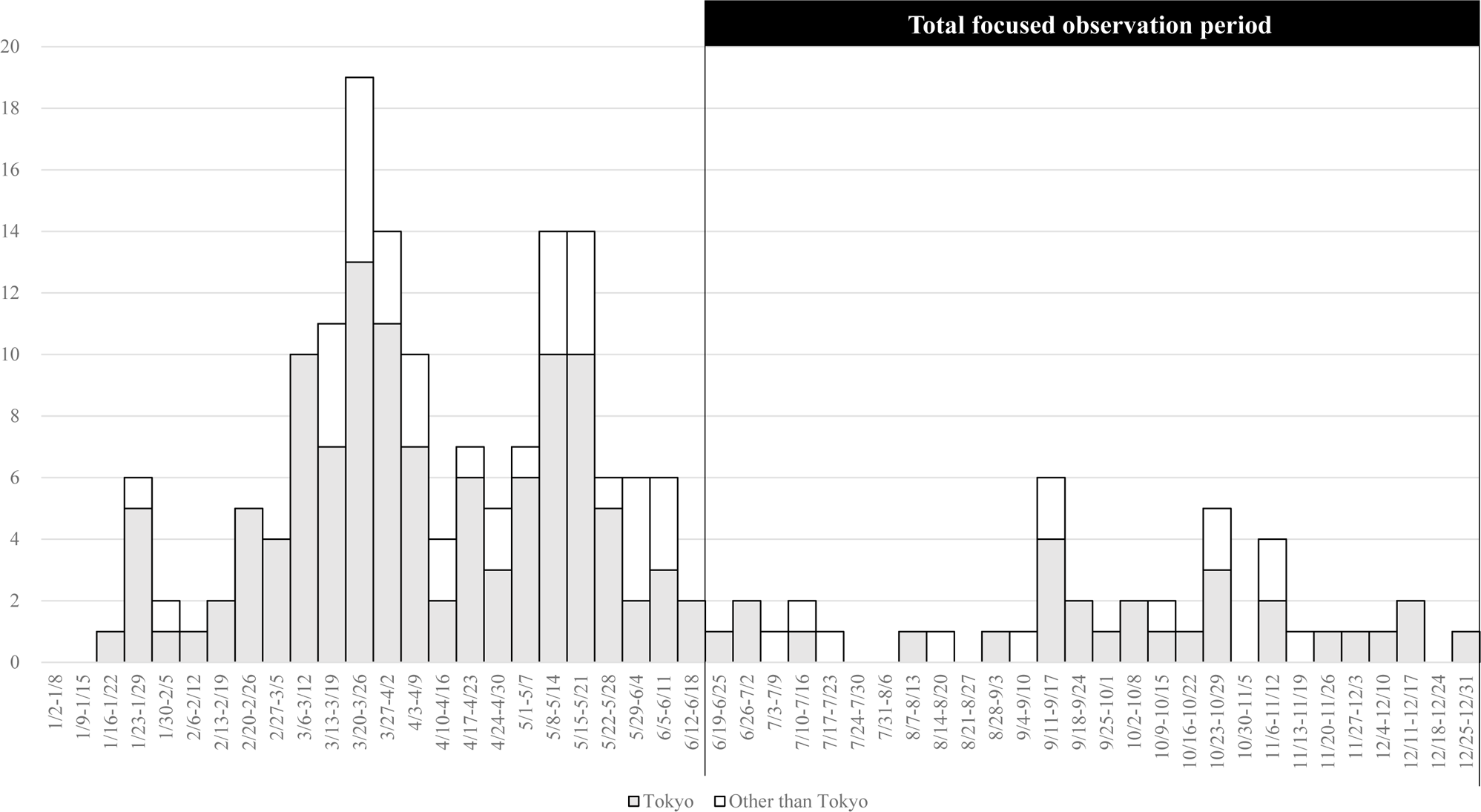
Number of reported mpox cases in Japan in 2023.

**Table S1.**
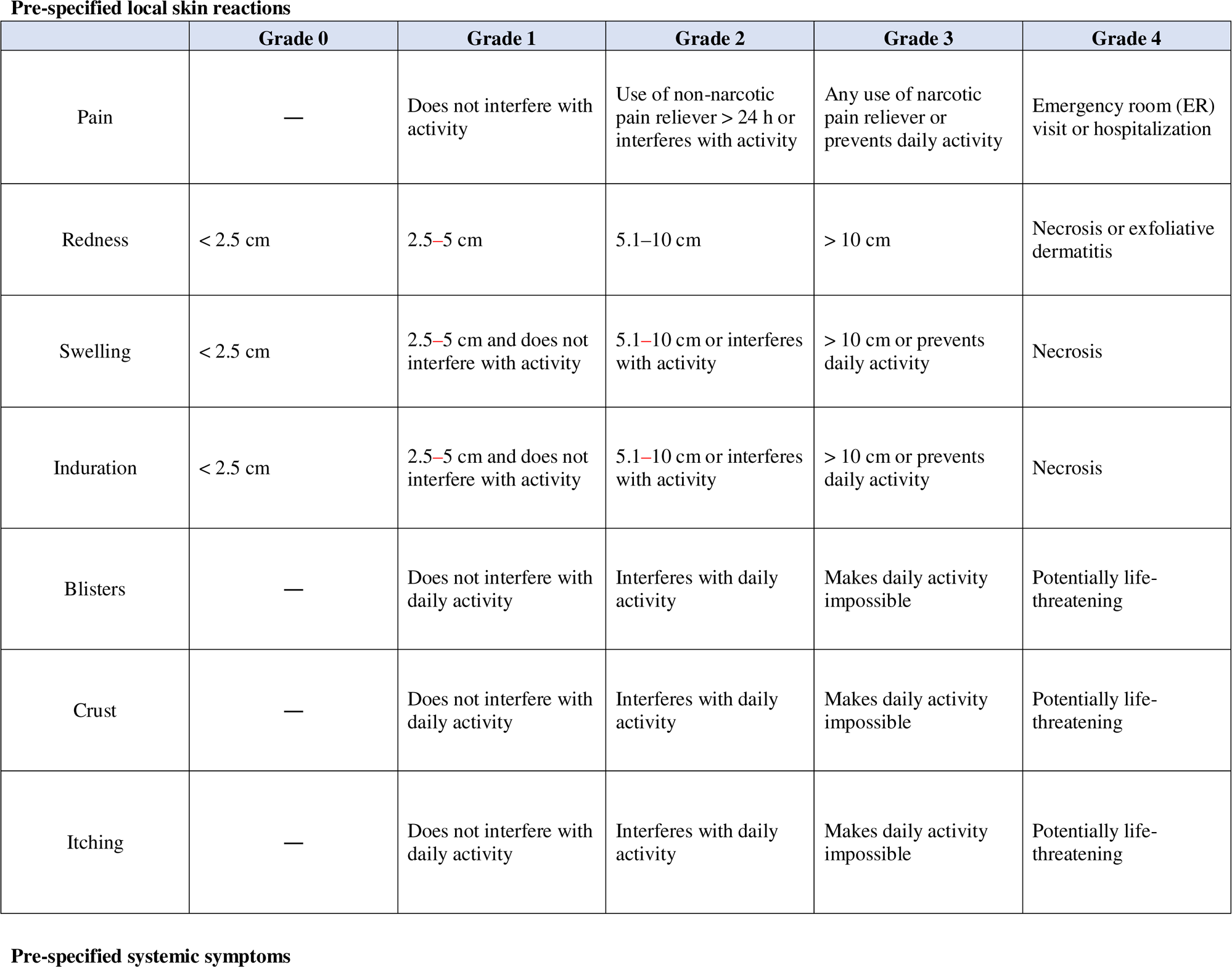
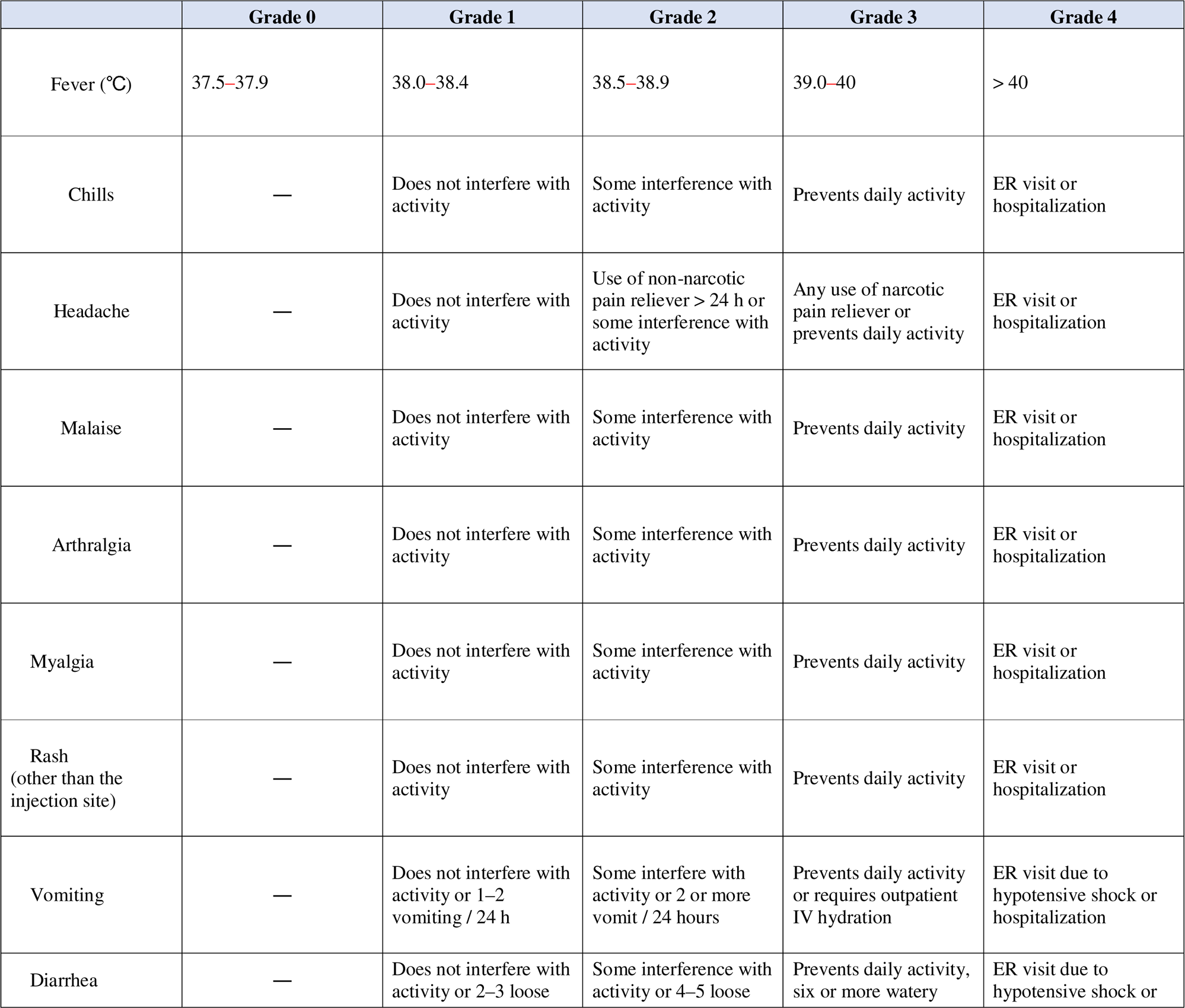
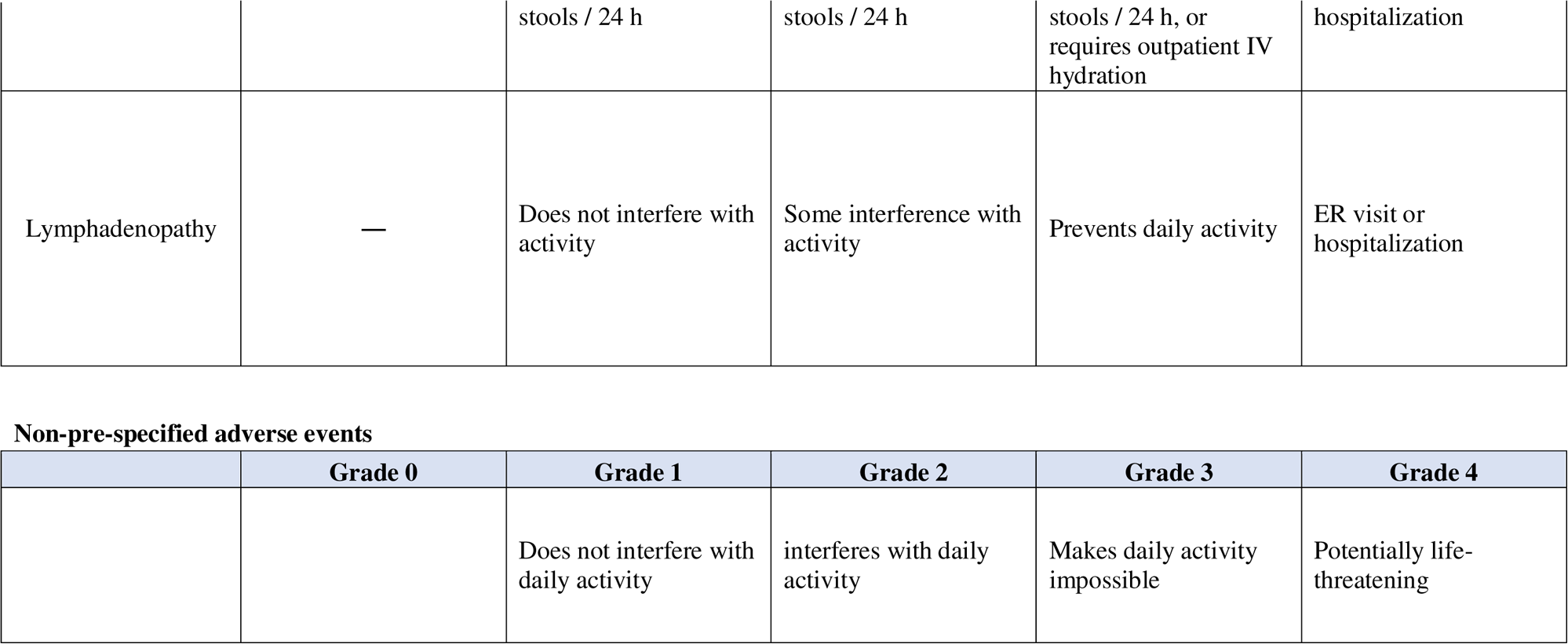
Grade of adverse events.

**Table S2.** Vaccine effectiveness for mpox development during entire observation period.

|  | N | Case | Mpox development |  | Vaccine effectiveness* |  |  |
| --- | --- | --- | --- | --- | --- | --- | --- |
|  |  |  | Observed<br>person-years | Incidence rate (100<br>person-years) | VE (%) | 95% CI | P-value** |
| M-ITT analysis population |  |  |  |  |  |  |  |
| Post-vaccination period | 1129 | 0 | 314.146 | 0.00 | - | - | - |
| Pre-vaccination period | 1129 | 0 | 193.706 | 0.00 |  |  |  |
| ITT analysis population |  |  |  |  |  |  |  |
| Post-vaccination period | 1135 | 0 | 314.858 | 0.00 | - | - | - |
| Pre-vaccination period | 1135 | 1 | 194.264 | 0.51 |  |  |  |

\*: $VE (\%) = (1-RR) \times 100$ RR (relative risk) estimated using modified Poisson regression. RR is synonymous with IRR (incidence rate ratio) in this study.
\*\* : one-sided test
M-ITT, modified Intention to Treat
VE, vaccine effectiveness
CI, confidence interval
RR, relative risk

**Table S3.**
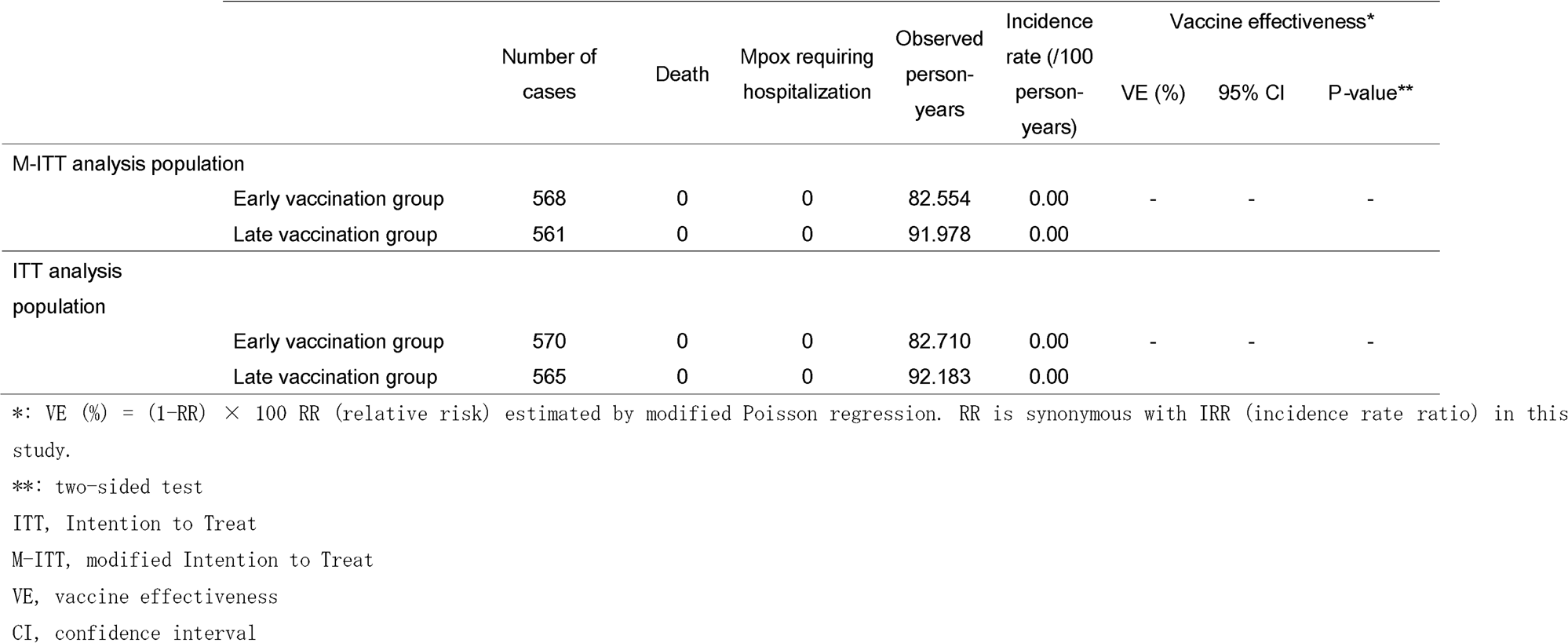
Vaccine effectiveness for severe mpox (death or hospitalization) during focused observation period.

|  |  | Number of cases | Death | Mpox requiring hospitalization | Observed person-years | Incidence rate (/100 person-years) | Vaccine effectiveness* |  |  |
| --- | --- | --- | --- | --- | --- | --- | --- | --- | --- |
|  |  |  |  |  |  |  | VE (%) | 95% CI | P-value** |
| M-ITT analysis population |  |  |  |  |  |  |  |  |  |
|  | Early vaccination group | 568 | 0 | 0 | 82.554 | 0.00 | - | - | - |
|  | Late vaccination group | 561 | 0 | 0 | 91.978 | 0.00 |  |  |  |
| ITT analysis population |  |  |  |  |  |  |  |  |  |
|  | Early vaccination group | 570 | 0 | 0 | 82.710 | 0.00 | - | - | - |
|  | Late vaccination group | 565 | 0 | 0 | 92.183 | 0.00 |  |  |  |

\*: $VE\ (\%) = (1-RR) \times 100$ RR (relative risk) estimated by modified Poisson regression. RR is synonymous with IRR (incidence rate ratio) in this study.
\*\*: two-sided test
ITT, Intention to Treat
M-ITT, modified Intention to Treat
VE, vaccine effectiveness
CI, confidence interval

**Table S4.** Take proporti.

|  | Number of cases | Take *sensu lato* |  | Take *sensu stricto*<br>(definition 1) |  | Take *sensu stricto*<br>(definition 2) |  |
| --- | --- | --- | --- | --- | --- | --- | --- |
|  |  | n (%) | 95% CI | n (%) | 95% CI | n (%) | 95% CI |
| Early vaccination group | 530 | 517 (97.5) | 95.8, 98.7 | 497 (93.8) | 91.4, 95.7 | 495 (93.4) | 90.9, 95.4 |
| Late vaccination group | 476 | 451 (94.7) | 92.3, 96.6 | 440 (92.4) | 89.7, 94.6 | 434 (91.2) | 88.3, 93.6 |

CI, confidence interval

